# Prevalence of Substance Use and related Behaviors among Tertiary Students: A Cross-sectional Survey in Yaoundé, Cameroon

**DOI:** 10.1101/2024.01.09.24301042

**Authors:** Michel Franck Edzamba, Fabrice Zobel Lekeumo Cheuyem, Adidja Amani, Tatiana Mossus

**Affiliations:** Department of Public Health, Faculty of Medicine and Biomedical Sciences, The University of Yaoundé I, Yaoundé, Cameroon

**Keywords:** substance use, university students, adolescent, youth behavior, Cameroon

## Abstract

**Background:** The use of psychoactive substances is a major global public health problem. People aged 15-24 years are more likely to abuse psychoactive substances than the general population. Substances use among youth, especially students in higher education is increasing rapidly worldwide. This study aimed to assess the prevalence of substances use among university students and to describe their behavioral profiles.

**Methods:** An institutional-based descriptive and cross-sectional study was conducted from September to October 2023 at the Yaoundé 1 University in Cameroon. A convenience non probabilistic sampling method were used to recruit consenting students. The data collectors were medical students who were trained for 2 days and given appropriate instructions before the survey. The data collected were reviewed and checked for completeness before being entered. All data were coded and entered into Microsoft Excel 2016. Descriptive statistics were performed using R Statistics 4.3.1. Qualitative data were collected from all participants through interviews.

**Results:** A total of 191 university students were enrolled in the study. The median age was 20 years. They were predominantly male (66.5%) and aged between 20-25. The prevalence of substance use was 66% for alcohol consumption, 33.5% for smoking habits and 26.7% for drug use. The main motivations for substance use were companionship, thrill-seeking and curiosity. More than a third of students were polysubstance users (35.1%). Most participants were aware of the academic consequence (85.9%) and almost all (93.7%) acknowledged the physical and psycho-social consequences of substance use.

**Conclusion:** There is a high rate of psychoactive substance use among university students. Therefore, effective campus-based counseling, peer education, and national surveillance systems that can monitor risky behaviors among university students should be implemented.

## Background

Psychoactive substances are those that produce pleasure for their users by altering perception and sensations, resulting in inner peace and satisfaction [1]. However, these pleasurable effects can be detrimental to an individual’s well-being, family relationships, social interactions, and professional pursuits [2]. Currently, an estimated 2 billion individuals worldwide consume alcohol, while 1.3 billion smoke cigarettes and 185 million take drugs. Each year, 5.4 million individuals die from smoking, and 76.3 million suffer from disease related alcohol use [3,4].

Alcohol is the most commonly consumed psychoactive substance worldwide. However, cannabis is the preferred illicit substance in Africa, while opiates are the most commonly used illicit substance in Europe and Asia, and cocaine predominates in South America [5]. People aged 15-24 years are more likely to abuse psychoactive substances than the general population [6,7]. In Africa, it was estimated that 8.4% of people aged 15 and 64 engaged in drug use in 2017 [8]. Furthermore, the projected total number of drug users on the continent is expected to increase by 38% between 2018 and 2030 [9].

Young adults are particularly vulnerable to experimentation, and the university setting presents inherent risks for substance-use related behaviors [10]. This, combined with rapid socioeconomic and cultural change and westernization in most sub-Saharan African countries, provides an enabling environment [11,12]. In addition, increased social contact with peers in the university community may influence initiation of substance use [13]. The transition from secondary school to university is characterized by considerable peer influence, and some students may be intellectually, emotionally and socially vulnerable throughout their years of study [14].

In Cameroon, according to the National Anti-Drug Committee, 21% of the population has used drugs. Of these, 60% are young adults aged of 20 and 25, some of whom are university students. In addition, more than 12 000 children under the age of 15 use narcotics and other psychotropic substances [8]. Students in higher education institutions most frequently use tobacco, alcohol, and opioids as psychoactive substances [12,15,16]. Alcohol and tobacco are considered “gateway” substances, because their use can lead to the consumption of so-called hard drugs such as heroin, cocaine, crystal methamphetamine, and ecstasy [8,17]. Data on the use of psychoactive substances among university students in Cameroon are scarce. This study aimed at assessing the prevalence of addictive substances use among university students and describe their behavioral profiles.

## Methods

### Study design, period and

An institutional-based descriptive and cross-sectional study was conducted from September to October 2023 at the Yaoundé 1 University in Cameroon. It was a mixed-methods study that included both quantitative and qualitative data.

### Setting

Yaoundé is the political capital of the Central Region of Cameroon. The population is estimated at 1.5 million and includes all of the country’s ethnic groups. The demographic structure is characterized by a very young population (people under the age of twenty make up 60% of the population) [18]. The University of Yaoundé I is a non-profit public institution of higher education institution located in the metropolis of Yaoundé. It educates between 10,000 and 15,000 students each year. The university has many faculties and professional training schools, the most important of which are in the fields of literature, fundamental sciences, medicine and education.

### Study participants & selection criteria

All students attending classes at the University of Yaoundé during the study period were eligible. All eligible students aged 18 years and above from the selected tertiary institutions, irrespective of their year of study and who gave their written informed consent were included.

### Sampling method

A convenience non probabilistic sampling method were used to recruit consenting students.

### Data collection tool and procedures

An interview was conducted by trained investigators to collect sociodemographic information and data on smoking habits (cigarettes, hookah), alcohol and drug use (marijuana, cocaine, tramadol, heroin, etc.). The sample population was recruited from students on campus. Prior to enrollment, potential participants were informed of the aims and objectives of the study before written informed consent was obtained.

### Data quality assurance

Data collectors were medical students who received 2 days of training and instructions prior to the survey. The data collected were reviewed and checked for completeness prior to data entry.

### Data processing and analysis

All data were coded and entered into Microsoft Excel 2016. Questionnaires were kept in a secure location accessible only to the principal investigator only. Descriptive statistics were performed using R Statistics 4.3.1.

Qualitative data were collected from all participants through interviews. Processing these data using content analysis techniques enabled the identification of suggestions for addressing substance use among students.

## Results

### Socio-demographic Profile

A total of 191 university students were enrolled in the study. The median age was 20 years. They were mostly male (66.5%) and aged 20-25. Students living with the family (67.5%) or residing outside (72.8%) the campus were the most represented (Table 1).

**Table 1.** Socio-demographic characteristic of study participants, Yaoundé, September 2023 (*n*=191)

| **Characteristic** | **Count(*n*)** | **Frequency (%)** |
| --- | --- | --- |
| **Sexe** |  |  |
| Female | 64 | 33.5 |
| Male | 127 | 66.5 |
| **Age group** |  |  |
| <20 | 71 | 37.2 |
| 20-25 | 87 | 45.5 |
| 25-30 | 23 | 12.0 |
| 30-35 | 7 | 3.7 |
| 35-43 | 3 | 1.6 |
| **Living situation** |  |  |
| Alone | 48 | 25.2 |
| Collocation | 14 | 7.3 |
| Family | 129 | 67.5 |
| **Residency inside campus** |  |  |
| Yes | 52 | 27.2 |
| No | 139 | 72.8 |
| **District of residence** |  |  |
| Biyem-Assi | 39 | 20.4 |
| Djoungolo | 8 | 4.2 |
| Efoulan | 100 | 52.4 |
| Mvog-Ada | 6 | 3.1 |
| Nkolbisson | 10 | 5.2 |
| Nkolndongo | 7 | 3.7 |
| Odza | 8 | 4.2 |
| Others | 13 | 6.8 |
| **Educational level** |  |  |
| Bachelor | 155 | 81.1 |
| Master | 29 | 15.2 |
| Doctorate | 7 | 3.7 |
| **Faculty & schools** |  |  |
| Literature | 25 | 13.1 |
| Medicine | 25 | 13.1 |
| Polytechnic | 9 | 4.7 |
| Sciences | 127 | 66.5 |
| Others | 5 | 2.6 |
| **Social charge** |  |  |
| Yes | 27 | 14.1 |
| No | 164 | 85.9 |

### Alcohol Use

Two-thirds of students reported that they usually drink alcohol (66.0%, 95% CI: 58.8-72.7%) with an average of 2.6 glasses of alcohol per occasion. More than half of the students reported drinking alcohol at least once a month (55.6%) and almost half said they felt the need to stop drinking alcohol (42.1%) (Table 2).

**Table 2.** Alcohol use related habits among Yaoundé university students, September 2023 (*n*= 126)

| Characteristic | Count (n) | Frequency (%) |
| --- | --- | --- |
| **Duration** |  |  |
| <6 months | 13 | 10.3 |
| 6-12 months | 19 | 15.1 |
| 1-5 years | 50 | 39.7 |
| 5 years + | 44 | 34.9 |
| **Frequency** |  |  |
| 2-3 times per week | 16 | 12.7 |
| At least 4 times per week | 13 | 10.3 |
| At least once a month | 70 | 55.6 |
| 2-4 times per month | 27 | 21.4 |
| **Feel the need to stop** |  |  |
| Yes | 53 | 42.1 |
| No | 73 | 57.9 |

Most respondents reported equally drinking beer, wine or whisky equally (43-46%). Traditionally produced alcohol was a less preferred type of alcohol (Figure 1).

**Figure 1.**
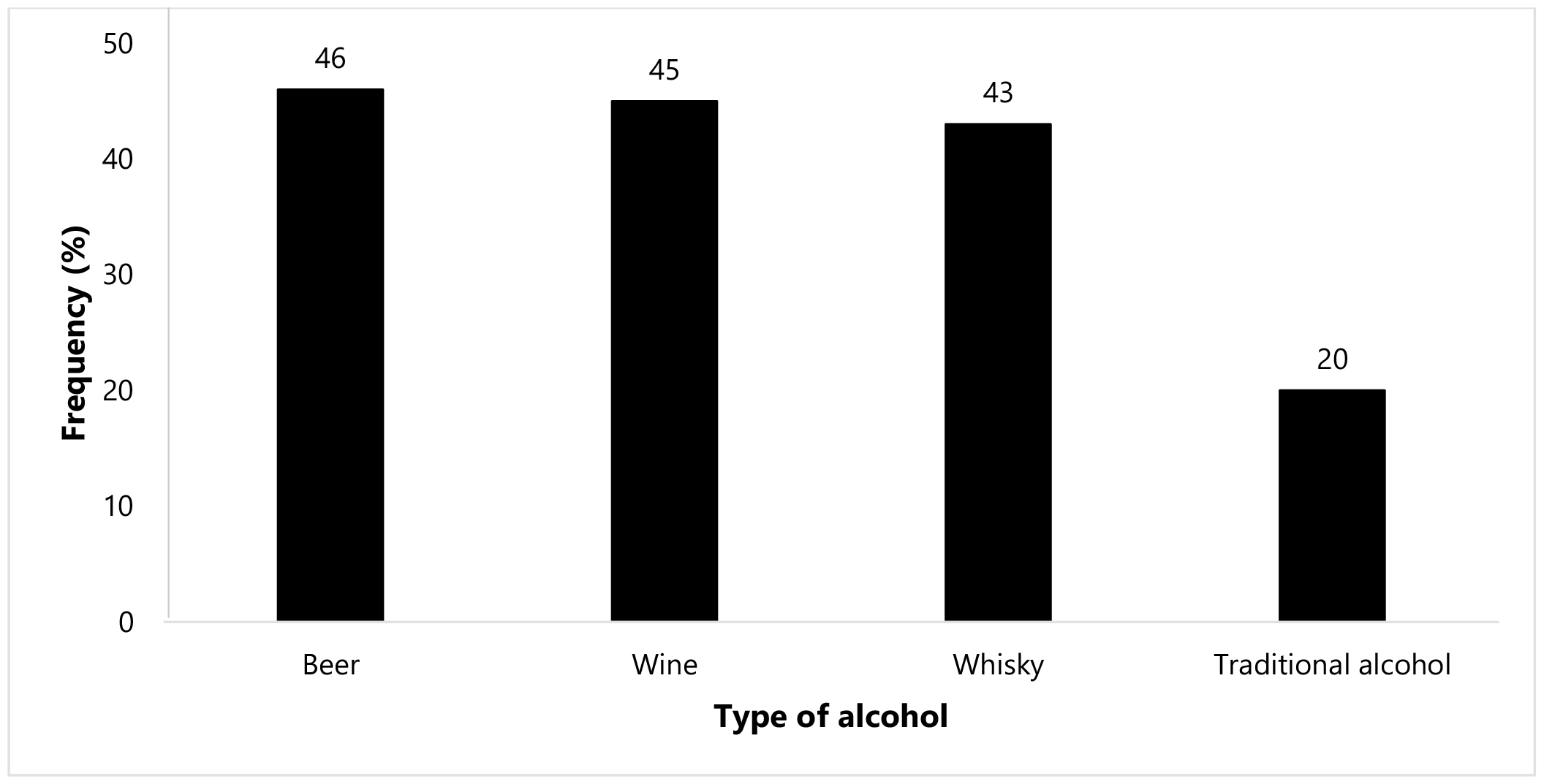
Type of alcohol usually consumed among Yaoundé university students, September 2023, (*n*=126)

Students mentioned companionship, emotion and trill-seeking as the mail reason motivating alcohol use. Almost a quarter of respondents reported no motivation for this behavior (23%) (Figure 2).

**Figure 2.**
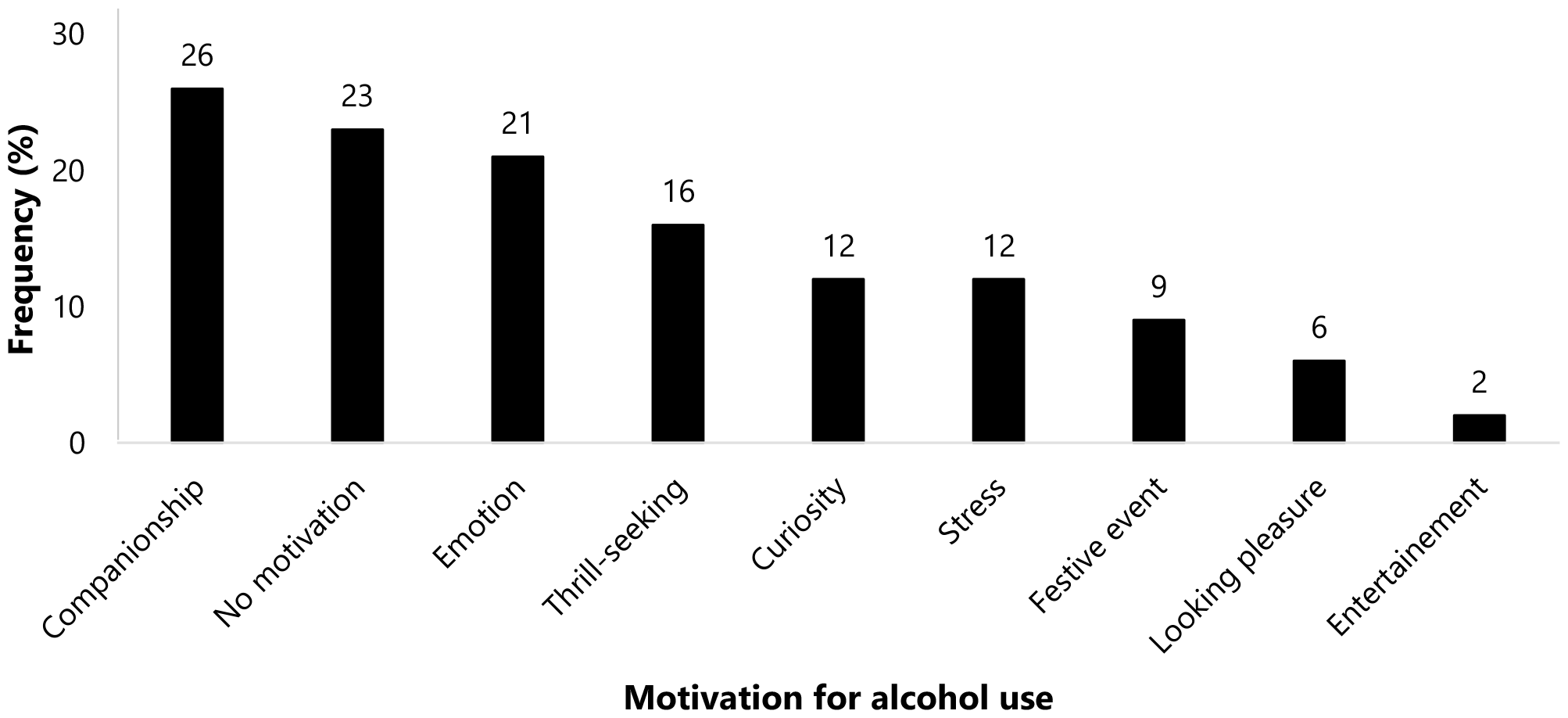
Factors motivating alcohol use among university students in Yaoundé, September 2022 (*n*=126)

**Figure 3:**
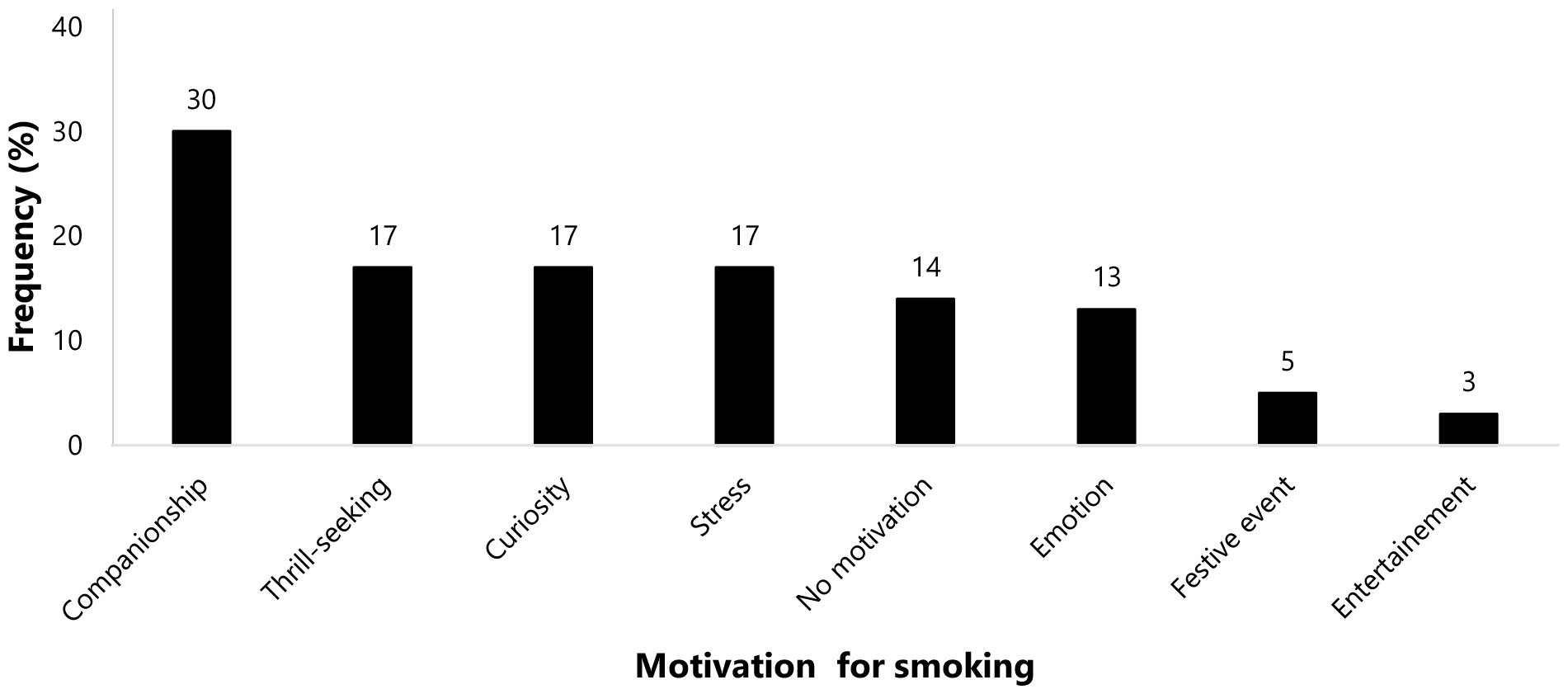
Factors motivating smoking among university student in Yaoundé, September 2022 (*n*=64)

### Smoking Patterns

The smoking prevalence among university students was 33.5% (95% CI: 26.8-40.7%). Most of participants reported smoking in the past 1-5 years (43.8%). Hookah (53.1%) was the most popular type of smoking among students. Cigarette users reported smoking an average of 6.2 cigarettes per day. Some two-thirds of respondents expressed the need to quit smoking (64.1%) (Table 3).

**Table 3.** Smoking habits among Yaoundé university students, September 2023 (*n*=64)

| Characteristic | Count (n) | Frequency (%) |
| --- | --- | --- |
| **Duration** |  |  |
| <6 months | 9 | 14.1 |
| 6-12 months | 18 | 28.1 |
| 1-5 years | 28 | 43.8 |
| >5 years | 9 | 14.1 |
| **Last smoking** |  |  |
| <6 months | 37 | 57.8 |
| 6-11 months | 11 | 17.2 |
| 12 months + | 16 | 25.0 |
| **Smoking type** |  |  |
| Cigarette | 15 | 23.4 |
| Hookah/Waterpipe | 34 | 53.1 |
| Both | 15 | 23.4 |
| **Need to smoke more** |  |  |
| Yes | 18 | 28.1 |
| No | 46 | 71.9 |
| **Need to stop** |  |  |
| Yes | 41 | 64.1 |
| No | 23 | 35.9 |

The most reported motivations for smoking included companionship (30%), thrill-seeking, curiosity and stress (17%).

### Drug Use Profile

The most reported known drug to university students included cocaine, tramadol and cannabis (Figure 4).

**Figure 4.**
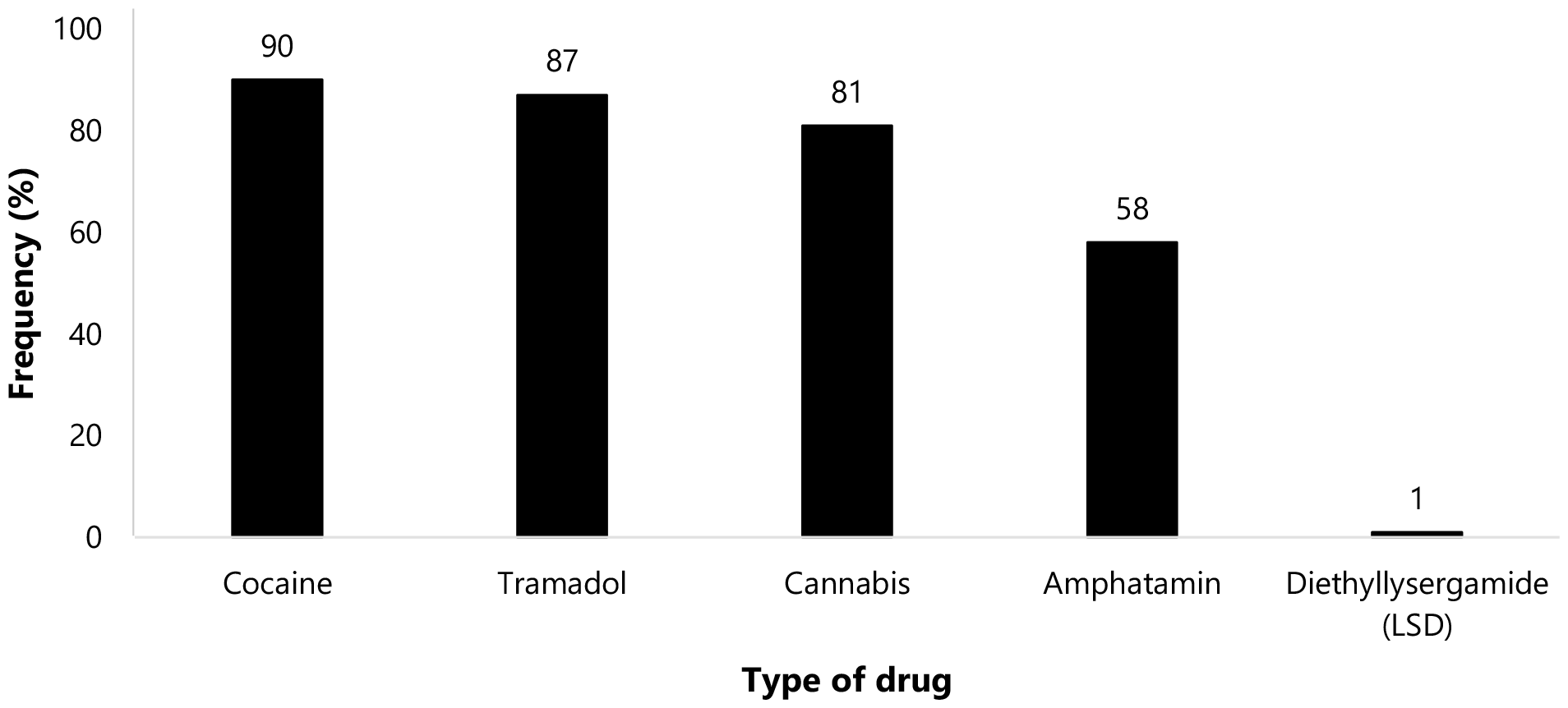
Drugs known to university students in Yaoundé, September 2023 (*n*=166)

The prevalence of drug use among students was 26.7% (95% CI: 20.6-33.6%). Almost a quarter of drug users reported having used the drug in the past week. More than two-thirds of respondents reported feeling the need to stop using the drug (Table 4).

**Table 4.** Drug use habits among university students in Yaoundé, September 2022 (*n*=51)

| Characteristic | Count (n) | Frequency (%) |
| --- | --- | --- |
| **Drug frequency** |  |  |
| Last week | 11 | 21.6 |
| Last month | 3 | 5.9 |
| Once or twice in your life, but in the last 6 months | 6 | 11.8 |
| Once or twice in your life, but not in the last 6 months | 31 | 60.8 |
| **Need use more** |  |  |
| Yes | 12 | 23.5 |
| No | 39 | 76.5 |
| **Need to stop** |  |  |
| Yes | 37 | 72.5 |
| No | 14 | 27.5 |

The most commonly reported motivation for using drugs was the company of close friends and relatives who are drug users (29%), curiosity (25%) and thrill-seeking (12%). However, some drug users expressed no peculiar motivation (20%) (Figure 5).

**Figure 5.**
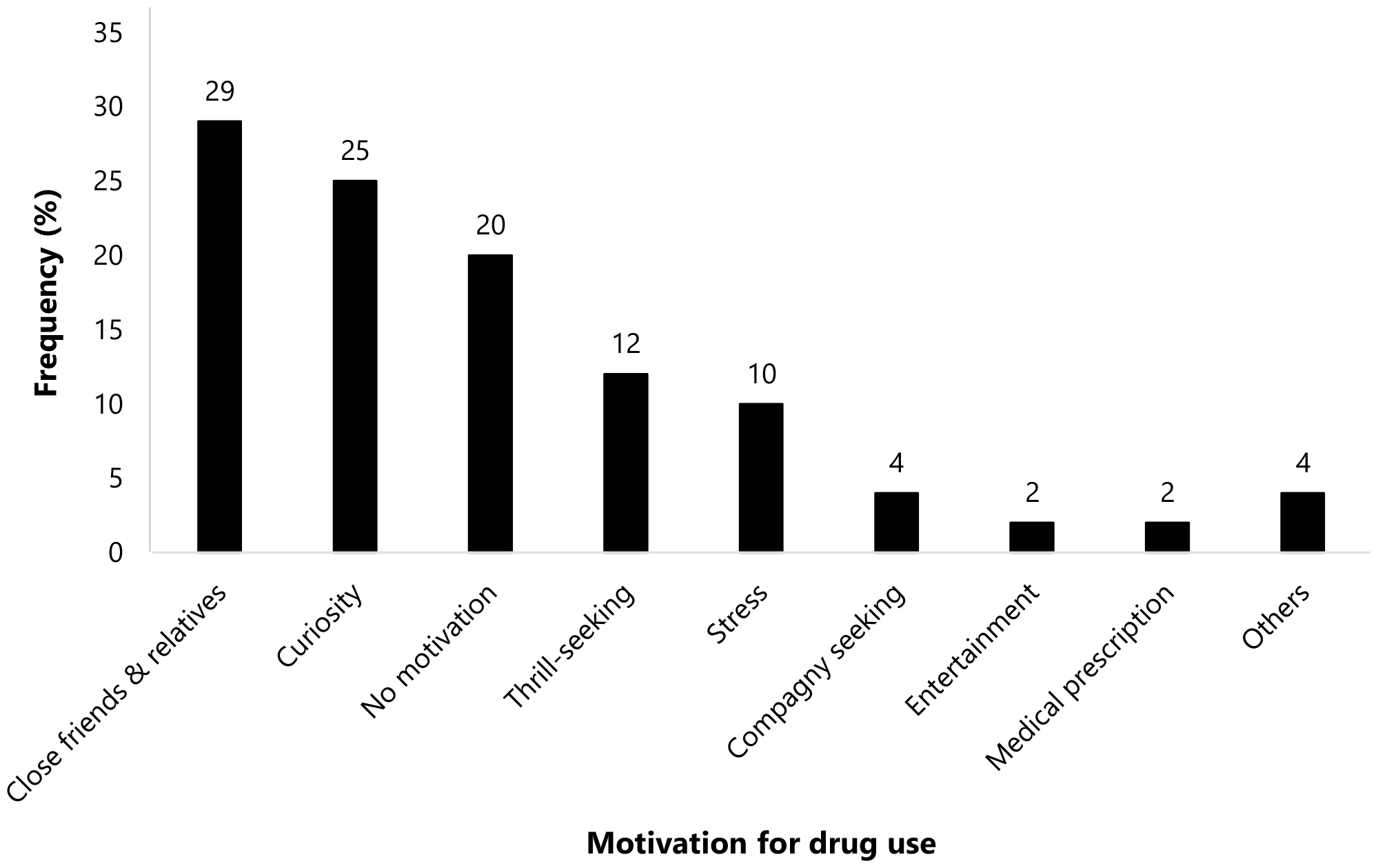
Factors motivating drug use among university students in Yaoundé, September 2023 (*n*=51)

More than one-third of students were polysubstance users (35.1%).

### Perception of Addictive Substances Use Consequences

Most participants were aware of the academic consequence (85.9%) and almost all (93.7%) acknowledged the physical and psycho-social consequences of substances use (alcohol, smoking, drug).

To the question on how to address the addictive substance use in the student milieu, most of them mentioned education and sensitization as the best method to be implemented.

To this regard, a participant suggested: « Seminars are needed to educate students, and the authorities must commit to taking measures to reduce consumption ». A Polysubstance user said: « Seminars sensitization on the disadvantages of alcohol and drugs on students ».

Other respondents mentioned the involvement of decision-makers: « The price of those substance i.e., alcohol and cigarettes should be increased » and a polysubstance user declared: « I think authority should reduce the rate of implementation of new bars ».

## Discussion

The use of alcohol, tobacco, and drugs is a social determinant that leads to the premature death of millions of people worldwide each year. Young adults are particularly affected by the global burden of disease related to substances use, as harmful habits are often initiated in this age group [3,4].

### Alcohol Use

The prevalence of alcohol use was 66%, and most students reported occasional use. This result corroborates findings from Ethiopia and Kenya [12,15]. However, this proportion was lower than that found in Buea (Cameroon), Nigeria and South Africa [1,8,19]. The difference in prevalence may be due to the larger sample size and the study period. Participants in this study were mostly living with their families, which may have influenced the consumption patterns. It is worth noting that the prevalence of alcohol use in this study was higher than that found in France (20.1%), Turkey (26.9%), and Myanmar (20.3%) [13,20,21]. Geographical differences in beliefs, religious and cultural practices, environmental factors, accessibility and availability of alcohol, urbanization, lifestyle, national laws and governmental and legal enforcement of alcohol laws may explain the observed differences in alcohol consumption [22].

Companionship, lack of motivation, and curiosity were the most commonly reported reasons for drinking. Previous studies have shown that adjustment to university environment, living away from family, low parental supervision and peer pressure (companionship) are predictive factors for alcohol use [21]. In fact, students who have friends who drink are seven times more likely to drink alcohol than those who do not. This finding confirms observations elsewhere in America [23].

### Smoking Patterns

The study found that 33.5% of the students smoked, more than half reported using hookahs (53.1%) and almost a quarter smoked either cigarettes (23.4%) or both (23.4%). These findings are similar to reports from Saudi Arabia [24,25]. These results suggest that cigarettes and water pipes are the preferred smoking devices among young smokers. This information could be emphasized in future health education campaigns.

The prevalence of cigarette smoking was 23.4%, which is comparable to the rates found in France, Myanmar and Ethiopia [12,20,21], but higher than those reported in other studies conducted in Nigeria [11,26]. However, it was lower than the rates found in a study conducted in Kenya (42.8%) [15]. The difference in study setting could explain these differences. It has been reported that students who have friends who smoke are five times more likely to smoke themselves. Several studies have examined the impact of peer influence on smoking behavior. Peer pressure is often cited as the primary reason for smoking initiation [27,28]. This study found that companionship, thrill seeking and curiosity were the main motivators for smoking. In addition, research has shown that 41% of smokers attempt to quit each year [29]. In this study, almost two-thirds of smokers (64.1%) were motivated to quit smoking, while only 28.1% expressed a desire to smoke more. This warranties the urgent need for university outreach strategies to identify students in need and provide them with temporary addiction specialists to address their desire to quit smoking.

### Drug Use

Almost all students reported that were familiar to various types of drugs. Of 126 participants, 26.7% said they had used drugs, and almost a quarter (21.6%) had done so in the week before the survey. This result shows that students have tried drugs less often than other substances. This can be explained by the fact that these drugs are not easily available to students, and that the possession and use of these drugs are punishable by the law of the land. In addition, it could be due either to underreporting due to the data collection method (interview) or to a lack of availability of these substances (which are certainly expensive). Nevertheless, this result was higher than findings from other reports [8,12,26]. Most of students reported being aware that substances use can affect their academic performance and be detrimental to their health corroborating observations elsewhere in Cameroon [8]. This warranties that more than two-thirds of the participants (72.5%) expressed a desire to stop using drugs.

### Polysubstance use and health conditions

More than a third of students were polysubstance users (35.1%). This result was higher than that found among adolescents in Vanuatu and Tonga (12.5%) [30]. The difference in the target population may explain this observation. On the other hand, report from Canada outlined of a higher prevalence of polysubstance use among youth (50%) [31]. Studies findings underline that the substance use is associated with high income and may explain the observed differences [8].

Drug use remains an area of concern because of the potentially dangerous short-term consequences including unintentional injury and death from traffic accidents [32].

People with addiction often have one or more co-occurring health problems, including lung or heart disease, stroke, cancer, or mental illness. Imaging scans, chest X-rays, and blood tests can show the damaging effects of long-term drug use throughout the body [33].

Tobacco smoke is now known to cause many cancers, methamphetamine can cause severe dental problems known as meth mouth, and opioids can lead to overdose and death. In addition, some drugs, such as inhalants, can damage or destroy nerve cells, either in the brain or in the peripheral nervous system [33].

Drug use can also increase the risk of infections such as HIV, hepatitis B and C through the sharing of injection equipment or unsafe practices such as sex without a condom [34,35]. Infection of the heart and its valves (endocarditis) and skin infections (cellulitis) can occur after exposure to bacteria through injecting drug use [33].

## Limitations

This is a descriptive cross-sectional study conducted before the start of the academic year, hence the small sample size. In addition, these results cannot be generalized to the entire population, and it is not possible to establish a causal relationship because substance use was determined by self-reported. It is important to note that 100% reliability cannot be guaranteed due to possible recall bias.

## Conclusions

The prevalence of lifetime use of psychoactive substances among university students is high. Alcohol is the most commonly used substance among students. This study underscores the need to urgently establish preventive measures including Information, Education and Communication activities on substances use among students. The university should have a unit responsible for organizing and developing strategies to prevent the harmful use of these substances among students. It is also recommended that government regulations regarding the production, sale, promotion, advertising, and consumption of alcohol and tobacco be strengthened. In addition, the extension of such a study to the national level should be explored in order to provide a more accurate picture of the problem in the student environment.

## Data Availability

All data produced in the present work are contained in the manuscript

## Abbreviations

## Declaration

### Authors’ Contribution

Study conception & design: MFE; Data collection: MFE and FZLC; Data analysis and interpretation: FZLC; Drafting of original manuscript: MFE and FZLC; Critical revision of the manuscript: MFE, FZLC, AA and TM; Final approval of the manuscript: All authors.

### Ethical Approval Statement

This study has received the authorization of the Faculty of Medicine and Biomedical Sciences (N°: 472/UY/FSMSB). Informed consent was obtained from participants prior to inclusion in the study. All methods were performed according to relevant guidelines and regulations.

### Consent for publication

Not applicable.

### Availability of data and materials

All data generated or analyzed during this study are included in this published article.

## Acknowledgements

Our gratitude goes to undergraduate students of the faculty of medicine who gave a support in the conduction of the study.

